# “They assume by having sex, they are a husband”: How Indian Female Sex Workers navigate relationship fluidity and sexual risk

**DOI:** 10.1101/2022.09.28.22280391

**Authors:** Subadra Panchanadeswaran, Shubha Chacko, Sel Hwahng, Guitele Rahill, Manisha Joshi, Ardra Manasi

## Abstract

There is limited evidence that highlights female sex workers’ (FSWs) agency in negotiating HIV risk in the context of multiple sexual relationships. Using a Gendered Vulnerability framework, this study employed a phenomenological approach to explore the FSWs’ active navigation of concurrent sexual relationships and their perceptions and assessments of HIV risk and sexual negotiation. Sixty (60) FSWs participated in the study. Data were collected through semi-structured interviews and focus group discussions. ATLAS.ti provided an interpretive framework for data analysis. Findings revealed that FSWs navigate a complex landscape of multiple sexual relationships that influenced their condom use decisions. Despite possessing accurate HIV knowledge, FSWs in this study indicated that multiple circumstances (e.g., economic constraints, damaging gender norms, merely paying transactional relationships with some clients, emotional attachments with others, gender-role bound relationships, and instances of sexual victimization) influenced their perceptions of power and their desires and perceived capacities for HIV risk and prevention decisions. A nuanced understanding of FSWs’ concurrent fluid relationships and the cultural contexts in which Indian FSWs work are vital when planning programs and policies for HIV prevention.

## Introduction

Globally, sex workers are at heightened risk for infection with the Human Immunodeficiency Virus (HIV), progressively developing Acquired Immunodeficiency Syndrome (AIDS) and disproportionately bearing the burden of the infection (Shannon et al., 2015). According to a UNAIDS report in 2014, HIV prevalence among sex workers is 12 times greater than among the general population. Many HIV prevention programs targeting female sex workers (FSWs) globally have focused on promoting personal behavior change, especially on emphasizing consistent and correct condom use with clients (Patterson et al., 2008; Shahmanesh et al., 2008; Stoebenau et al., 2009) with some success and have revealed that FSWs possess the desire and willingness to negotiate condom use with paying clients. However, past research also points to FSWs’ reluctance to use condoms with non-paying intimate or romantic partners, typically due to their perceptions of powerlessness within those relationships, their fear of partner violence, and their varied subjective assessments of the relationship’s meaning and of the risks that non-condom use present in those relationships (Deering et al., 2011; Fitzgerald-Husek et al., 2011; Jie et al., 2012; Murray et al., 2007; Ngugi et al., 2012; Panchanadeswaran, et al., 2008, 2010; Rosenthal & Oanh, 2006).

FSWs’ reticence to use condoms in intimate relationships is not unique to them, since their non-sex worker counterparts are influenced in that regard by relationship power, dominance, dependence on partners for intimacy and other needs, which are key determinants of their sexual behaviors (Drigotas & Rusbult, 1992; Pulerwitz, et al., 2000; VanderDrift, et al., 2013). Moreover, client type, relationships with non-paying partners, perceptions of HIV/STI risk, and economic disadvantages contribute to the lack of consistency in FSWs’ negotiation of condom use (Fehrenbacher, et al., 2016). Additionally, the widespread criminalization and stigmatization of sex work often consigns FSWs to covert engagement with their partners and to recurrent victimization by pimps, law enforcement personnel, and gang members (Erausquin et al., 2011; Karandikar & Prospero, 2010; Odinokova et al., 2014). Thus, FSWs often navigate multiple sexual relationships in which multiple factors influence their perceptions of how much power they can potentially wield, their subjective assessment of risk, and their strategies for staying safe. Nevertheless, there is limited acknowledgement of the complexity of the spectrum of sexual relationships that FSWs navigate on a regular basis and sparse evidence that highlights FSWs’ agency in negotiating HIV risk in the context of such multiple sexual relationships. To address this gap in the literature, this qualitative study explored how street and home-based FSWs in India assess their risk for HIV/AIDS within multiple sexual partnerships and negotiate condom use with concurrent (paying and non-paying) sexual partners. By shedding light on the complexity of FSWs’ fluid relationships, this study challenges dominant discourses of viewing sex workers as mere vectors of disease and highlights FSWs’ strategic and agentic decision-making to ensure their safety and well-being.

## Literature Review

### Sex Work and the Indian Landscape

Both historically and in contemporary India, sex work has been viewed through a colonial and moralistic lens, resulting in policies that have been largely punitive and discriminatory (Kotiswaran, 2001). While sex workers are criminalized on one hand, even public health interventions that seek to offer them health services can often reduce them to their sexual behaviors, without acknowledging how they navigate those relationships, or their relative autonomy or agency within those relationships (Vijayakumar, 2021). Researchers in the past have pointed to the problematic conflation of sex work with the issue trafficking of women and the resulting monolithic portrayals of female sex workers as hapless victims without agency and undeserving of protection from discrimination (Ahmed & Seshu, 2012; Kaiwar & Gothoskar, 2014; Kotiswaran, 2011). Recent evidence contests notions of FSWs as involuntary victims of sex trade and emphasizes their active role in choosing sex work not only as a viable economic opportunity in addition to recognizing sex work as an empowering opportunity to make autonomous choices about their work and bodies (Azhar, et al., 2020; Karandikar, et al., 2022). Central to the recent scientific literature concerning FSWs’ condom use behavior is the concept of relationship fluidity.

### Relationship Fluidity

The concept of relationship fluidity denotes that for sex workers, “clients and nonpaying partners may not always represent distinct types of relationships. Rather, these are fluid relationships, distinguished from one another through the development of affective ties over time” (Stoebenau et al., 2009, p.811). Relationship fluidity can also loosely pertain to relationships of emotional intimacy that are accompanied by economic expediency. Fluid relationships can heighten FSWs’ vulnerability to HIV when they forego condom use with intimate partners with whom they deem condom use as inappropriate and consign condom use to sexual liaisons with only paying clients (Stoebenau et al., 2009). Accordingly, Stoebenau and colleagues (2009) have argued for a more nuanced understanding between paying clients and non-paying partners, cautioning against treating sexual relationships of FSWs as “belonging to separate, mutually exclusive categories” (p. 11), and called for an examination of the meaning that FSWs ascribe to their various relationships in association with using condoms to prevent HIV. Given the scant literature on a comprehensive understanding of sex workers’ subjective assessments of HIV risk in the context of concurrent sexual partnerships, this study attempted to shed light on FSWs’ decision making in the context of relationships with various partners.

Exploring relationship fluidity and HIV risk among FSWs in Karnataka State in India is important for several reasons. First, national estimates in India from 2019 reveal that there are 2.349 million individuals living with HIV/AIDS in the country, which translates to 0.22% prevalence among adults; and the prevalence of HIV/AIDS among FSWs ranges between 0.14-2.12% while the prevalence in Karnataka is 0.63% (National AIDS Control Organization & ICMR-NIMS, 2020). Second, since the start of the epidemic, Karnataka has been identified as one of the high-risk states where the majority of street and home-based FSWs are based and possess limited education and few marketable skills and face dire economic hardships, all of which influence many women’s entry into sex work (Panchanadeswaran et al., 2008; 2010) and thus it is imperative to examine those FSWs who are most economically vulnerable. Third, the socio-cultural context of FSWs in India is such that involvement in a non-paying heterosexual partnership often provides a dominant anchor and affirms prescribed gender norms of Indian society for FSWs (Deering et al., 2015; Karandikar & Prospero, 2010; Panchanadeswaran et al., 2008). Thus, the desire for a stable non-paying male partner may be as much about social viability as it is about economic security.

This study addresses the literature gap on the diverse contexts of sex work and complex sexual partnerships and condom use decisions holistically within FSWs’ various sexual partnerships in India. We acknowledge that sex workers vary in identity, including cisgender, transgender, and gender-non-conforming, heterosexual and homosexual, youth and adult sex workers, and they operate in diverse settings around the world. This study focuses on cisgender heterosexual female sex workers. We also recognize that cisgender heterosexual FSWs are not unique in their experiences of navigating a complex landscape of sexual partnerships and HIV risk, and that non-cisgender sex workers experience similar risks. However, experiences of non-cisgender women such as trans women who are FSWs are beyond the scope of the present work. The purpose of this study was to explore how cisgender heterosexual FSWs in India handle multiple and fluid sexual partnerships, what meanings they ascribe to these partnerships, their desires and their perceived power to negotiate safer sex in and across those relationships, and factors that may increase their HIV vulnerability. Specifically, we sought to elicit FSWs’ subjective, lived experiences of traversing diverse sexual partnerships and to obtain first-hand knowledge of how FSWs exercise their personal agency in the assessment of the nature of the sexual partnership. Further, we aimed to understand FSWs’ evaluation and tolerance of personal and biological (e.g., HIV) risk in the contexts of their various partnerships.

## Conceptual Framework

### Gendered Vulnerability

We used Gendered Vulnerability (Dutton et al., 2006) as a framework to explore how FSWs in India navigate multiple and fluid sexual partnerships, the different meanings they ascribe to these partnerships, their desire and perceived capacity to negotiate for safer sex in and across those relationships, and factors that may increase their HIV vulnerability. Gendered Vulnerability describes “[the] emotional, cognitive, behavioral, social, or physiological conditions that reduce [in the present case, a FSW’s] capacity or likelihood of resistance to coercion…” (Dutton et al., 2006, pp. 1-50). Gendered Vulnerability especially affects poor women around the globe who are socialized to accept male and societal domination and is a main reason why women and their children bear a disproportionate burden of extreme poverty, and in the present case, may be relegated to sex work (Jaggar, 2009). The Gendered Vulnerability framework may help explain how FSWs in the present study navigate multiple and fluid sexual partnerships. This study was also informed by the concept of “subjective bargaining power” of sex workers in India proposed by Hui (2017; p. 50). Specifically, Hui maintains that sex workers’ subjective assessment of the extent of control they have over structural, occupational, and personal constraints influences their individual capacity and power to negotiate or bargain for safer sex, which ultimately affects their well-being (Hui, 2017). The application of the concept of subjective bargaining power to this study is in its acknowledgement of the importance of FSWs’ subjective assessments based on their individual lived experiences.

## Methods

### Research Design

A phenomenological approach is used when the researcher is interested in obtaining the inside or emic perspective of individuals who possess experience about the phenomenon of study and when individuals’ unique insights into a phenomenon are desired. In the present work, a phenomenological approach was employed to better understand the complex nuances underlying various sexual partnerships for FSWs in India and to document the interpersonal, social, and cultural factors that influence their definition and use of safer sex behaviors. This approach is consistent with the theoretical framework comprising gendered vulnerability and the concept of subjective bargaining power, as it facilitated the empowerment of the participants as key informants whose emic perspectives were both valued and essential to this study, and, enabled researchers to draw on FSW’s lived experiences of negotiating condom use in India, in their own words.

### Participant Recruitment, Sample, and Sampling Procedures

Consistent with our objective of obtaining FSWs’ first-hand perspectives negotiating safe sex with various partners, we used a purposive sampling method, i.e., venue-based sampling. The venues included a key partner (a community-based organization-CBO) in an urban area and their affiliates in four rural areas. We recruited cisgender women who self-identified as sex workers. These women were recruited with the support of the bENGlocal CBO, a long-time research collaborator of the first and second authors. The collaborating CBO identifies as a workers’ union that has combined the approaches of a labor model as well as sex worker collectivization in addition to the ideas from the women’s and sexual minority movements (Panchanadeswaran, et al., 2016). The lead researcher (first author) explained the study goals in informal meetings with key leaders within the partner CBO and with community leaders who work closely with the partner CBO. Subsequently, information about the study was disseminated at the CBO’s community outreach meetings in the urban and four rural areas. Two professionally trained social workers served as research associates who assisted the lead researcher in participant recruitment and data collection. Interested FSWs who volunteered to participate were given the contact information of the research associates. Interested participants were screened for eligibility based on the following criteria: (1) aged 18 years or older, (2) had either exchanged sex for cash/kind in the previous three months, (3) solicited clients on streets and/or public venues such as cinema halls, bus terminals, railway stations, hotels/lodges and/or independently through brokers or through informal social networks, and/or (4) provided sexual services in a public place and/or in their own homes or in others’ homes. Our University’s Institutional Review Board approved the study. Our final sample comprised 60 FSWs.

#### Data Collection Methods

Data were collected through semi-structured, in-depth interviews (IDIs) (n=25) and five focus group discussions (FGDs) (n=35). Participants chose to participate in either of the modalities based on their convenience/preference. IDI participants roughly equally distributed between both rural (14) and urban (11) locations. One FGD was conducted with sex workers in the urban location (n = 6) and four were held with those in rural areas (n = 6,7,10,6). All the interviews and FGDs were conducted in the local language, Kannada. Additionally, the interviewer (first author or a research associate) also administered a short socio-demographic survey to all respondents. The researchers (first author or second author) reviewed the informed consent process with each participant, explained the study goals, and elicited both written and verbal informed consent in addition to explicit verbal consent for audio-recording. Participants were assured of confidentiality and were offered the opportunity to use pseudonyms as an additional step to protect their identity. Data were collected in private office rooms of the local collaborating partners at the indicated venues. In-depth interviews lasted between 45 minutes to an hour while FGDs lasted about 90 minutes. Interview guides and the short survey were piloted, revised, and finalized based on the feedback received from five community gatekeepers. In keeping with the local collaborating organizations’ protocol, participant compensations included travel reimbursement and a meal.

### Data Analysis

All interviews and focus group discussions were first translated and transcribed verbatim into English by three experienced local transcribers with a history of having worked with the sex worker population in Karnataka. The transcripts were then uploaded as Primary Documents into the hermeneutic framework of ATLAS.ti® 7.0 (Muhr, 2013), a computer assisted data analysis software. Atlas.ti has been used successfully by the authors to analyze FG transcripts of the lived experience of female victims of non-partner sexual violence in Haiti (Joshi et al., 2014) and to analyze data collected from sex workers in Mumbai and Bengaluru cities in India (Panchanadeswaran et al., 2017; Panchanadeswaran et al., 2016). ATLAS.ti® has also been a useful framework for analyzing qualitative data in other health and social science research and in fields outside of social science research (Summer et al., 2017).

Prior to open coding, the first and third authors read the transcripts multiple times. We followed the recommendations by Campbell, et al. (2013) to ensure rigor and inter-coder agreement in the data analysis procedures. Based on prior literature review and the study goals, the researchers developed a broad coding plan in line with the study goals. During open coding, the first step, both researchers read the transcripts line by line and independently coded a few sample transcripts to identify meanings pertaining to the study’s objectives. Sections of transcripts were assigned broad labels such as ‘unknown client’, ‘intimate partner’, ‘caring client’, ‘police violence’, ‘gang member violence’, ‘condom use negotiation’, ‘HIV risk’ etc. Subsequently, quotations were extracted from these sections of text. Special attention was paid to participants’ emic perspective of risk, agency, and safety. Analysis was not restricted to apriori codes but included insights that were not anticipated and allowed for emergent codes to be incorporated into the data analysis. For example, findings revealed insight into the complexity of entryways into sex work and other factors related to FSWs’ lived experiences in India which set the background for the initial type of transactional relationships with clients. Subsequently, the researchers reviewed the open codes, discussed the specific codes that could be merged for axial coding. and specific categories were developed using data reduction methods (Creswell, 1998). In the final step, based on the recommendations of Glaser and Strauss (1967), between and within comparison of categories were undertaken, categories merged resulting in the emergence of broad themes including different types of client relationships, intimate partners and other sexual partners, subjective HIV risk perceptions, perceived power, and decisions regarding condom use.

#### Methods to Ensure Rigor

The researchers employed several strategies to ensure rigor, i.e.,trustworthiness as recommended by Creswell (2007) and Berger (2015). As mentioned earlier, this study used feminist research methodologies that were grounded in capturing the respondents’ insights from their own standpoints in a dialogic conversational format (Haney, 2002). Managing reflexivity in this study was achieved in that all data were collected by the first and the second authors who, as middle-class urban women were mindful of how their social locations and identities affected the research process and have been transparent about these dynamics in their long-standing partnership with the local CBO. One distinct advantage was that both researchers spoke the local language and the second author, and the research associates were social workers/activists with over two decades of work experience with significant familiarity with the sex worker community. After each interview/FGD, the researchers conducted peer debriefing to discuss any emergent issues that could impact the research process. To establish credibility of the analytic process, the researchers maintained a written audit trail to document all decisions related to the research and analytic process. Peer review and external audit was through the presentation of a draft of the findings to the key stakeholders including the main partner CBO and two other collaborating NGOs as well as member checking (confirming with sex worker participants that the findings represent their experiences and views) with sex worker participants. All stakeholders agreed that the findings reflected their knowledge of the phenomena explored and were consistent with the experiences of the street and home-based FSWs in the geographical locations where the study was conducted.

## Results

### Participant Profiles

Participants in the current study were young (Mean 30 years, S.D. 7.75). The mean years of formal education was 6.33 years (SD. 3.8). A significant proportion (63.3%) belonged to either Dalit/Other Backward Caste, while 28.3% reported belonging to the dominant/land owning caste and five women did not report caste identification and self-identified as Christians/Muslims; 90% self-identified as Hindus. Most reported being separated/divorced (51.6%), widowed (10%), and about 21.6% reported being ‘married’ or cohabiting with partners in ‘marriage-like’ relationships. Most (82%) participants reported having at least one child (Mean 1.55, *SD* 1.02). The majority of FSWs in the current study (63.3%) reported being engaged in other jobs other than sex work to make ends meet, and for over half (56.7%) of the respondents, sex work was the primary source of income. Sixty percent of study participants comprised FSWs who resided in rural areas and 40% were from urban areas. Most of the sample (73.3%) had entered the sex work as young adults between the ages of 18-25 years (Mean 23 years) and reported engaging in sex work for over half of every month (Mean 15.6 days/month). FSWs met an average of 4.2 clients each week. Most (68.7%) reported streets/fields/farms as the primary site of sexual services while nearly a third (31.7%) reported providing services primarily in rented homes, lodges, and hotel rooms, but these were not mutually exclusive.

### Complex Pathways of Entry into Sex Work

In the present study, economic hardships were the main pathway of 85% FSWs’ entry to sex work. Much of this economic hardship stemmed from their attempts to escape abusive or neglectful marital relationships, in which they perceived no option other than sex work to earn money to support themselves and their children other than sex work:

> …. everything (after marriage) was good for one month. Then he started doubting me and accusing me and also beating me…He used to call me a bitch and ask with how many men I had relations with….he hurt me a lot. He said you are *dagaar* (an immoral woman, a prostitute), and told me, ‘Hey bitch, leave my home.’ [IDI: FSW between 20-24 years]

Relentless cycles of violence, emotional abuse, and suspicions of infidelity by husbands also compelled FSWs in the present study to voluntarily terminate their marriages, which left them with no economic support:

> He (husband) started abusing me, and he told me that the father of the child was somebody else! I became fed up with his abusive talk. I concluded that he doesn’t need me anymore, so there was absolutely no point in living with him, hence, I had to separate from him and find a way to support myself also. [IDI: FSW between 20-24 years]

Failed marriages owing to husbands’ alcoholism or extra-marital affairs were also commonly cited as reasons for women to opt for sex work to fend for their families:

> (I have) two children…. I need at least 1000 rupees ($15) a month for household expenses. I can’t rely on my husband because his earnings are just enough for his booze and merry making. It is not as if I can sit at home and run the house with anything from his earnings. He is also having many affairs with girls, so much of his earnings go to that. So, I had to find a way to make money to feed my children. [IDI: FSWbetween 20-24 years]

Most FSWs in the current study spoke at length about the economic unviability of their past or current history as employees in the informal sector and the influence of peers in their social networks on their consideration of sex work as a viable form of livelihood:

> I learned about sex work from women who worked at a factory where I used to stitch gunny bags…before that I was working in a garment factory…for 8 hours, I had to stand and stitch. My body could not take it anymore. An aunt of mine told me all about it (sex work) and convinced me. After a lot of work, in the factory, at the most, you may earn 2000-3000 rupees ($29.4-$44) per month, whereas, in this profession, so much amount can be earned from a single client itself. [IDI: FSW between 35-39 years]

The above evidence provides an understanding of the complexity of entry into sex work as a context for FSWs’ decision-making about condom use power and the perceived risks that correspond to each sexual partnership that they navigate continually. Participant narrations revealed their ongoing efforts to juggle multiple sexual partnerships as depicted in Figure 1 below:

**Figure 1:**
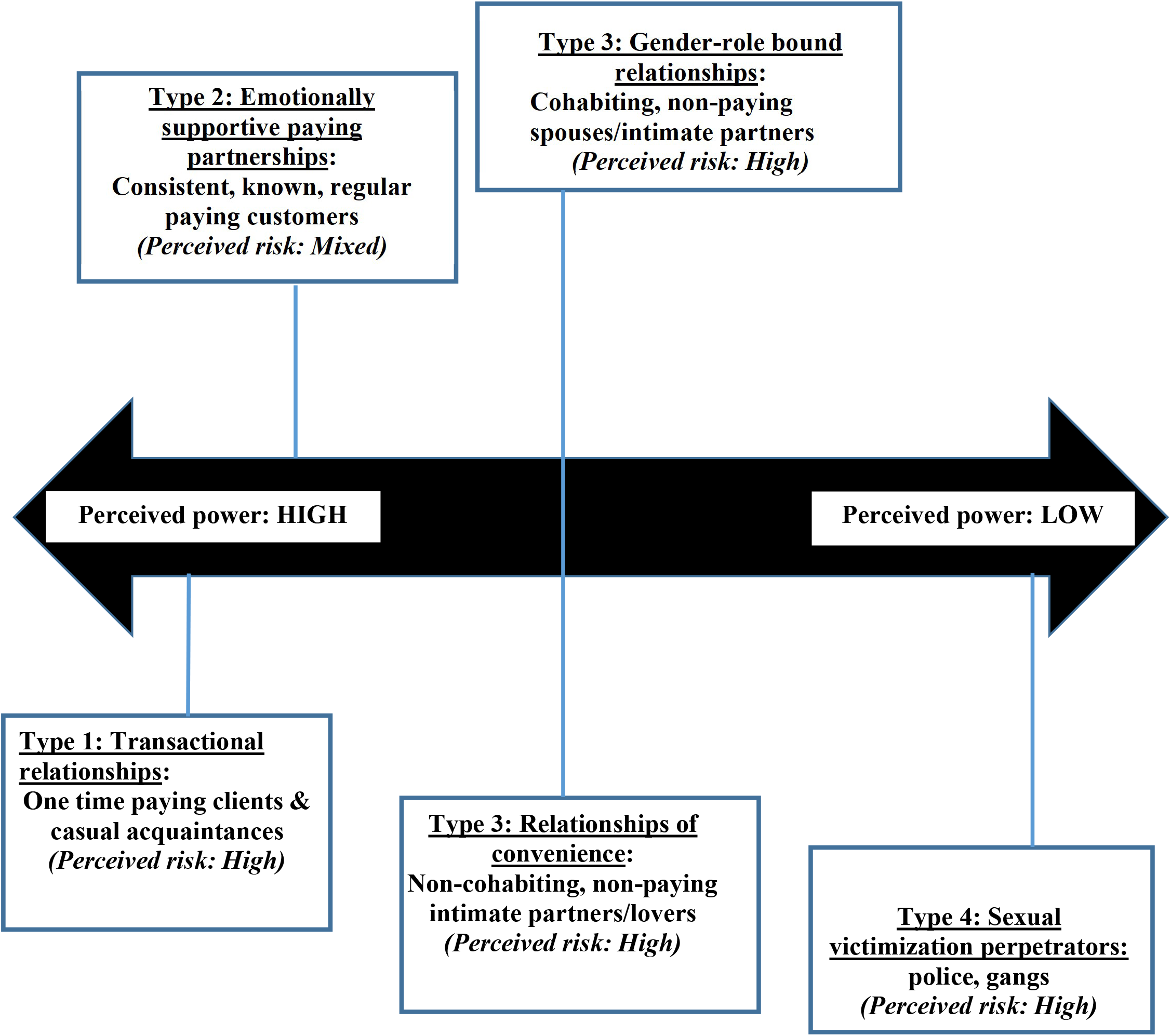
Fluidity of sexual partners, subjective assessment of sexual risk, and condom use on a continuum of female sex workers’ perceptions of power to negotiate safe sex

#### Fluidity of sexual partnerships, subjective perceptions of risk and FSWs’ perceived power over condom use negotiation

As depicted in Figure 1, participants’ narrations in the current study revealed the diversity of sexual partnerships that women navigated on a regular basis; these relationships range from purely transactional, paid relationships with unfamiliar clients, to sexual partnerships with seemingly caring clients who provide them with emotional and social support, to relationships of convenience with partners for purposes of gender stability and adherence to heteronormative social norms, through exploitative sexual encounters often marked by sexual coercion.

Interspersed with these relationships were traditional gender-role bound intimate or marital relationships. FSWs’ desire and perceived capacity to use condoms in sexual encounters with these various partner types were largely dictated by the nature of the sexual partnerships.

##### Transactional relationships: Type 1

The most common and obvious type of sexual partnerships that participants described were those with unknown, often first-time, paying clients. Without exception, all study participants spoke of business-like, un-entangled relationships with these unfamiliar clients. The transactional nature of the relationships with such clients appeared to place FSWs in relative positions of control and power. Significantly, respondents’ accounts underscored their perceptions of high risk for HIV from newer and unfamiliar clients, which made the reported condom use in those contexts almost unanimous. Most participants spoke of using a proactive, often strident conditional approach to condom use with one-time paying clients, as evidenced by the following quotes:

> I’ll hand them (condoms) over to the lodge people and instruct them to give it to every customer –even if they say no. (I say), ‘If not, don’t send them’. The lodge people charge 50 rupees ($0.77) per condom. The customer has to pay for it. Besides, the lodge people provide us security support. If the customer doesn’t pay us and tries to create unnecessary scenes, they handle such customers and make them pay. Without condoms I don’t have contact (sex) with customers, even if I have to starve… [IDI: FSW between 20-24 years] Madam, even if the client gives me 10,000 rupees ($135), I do not agree if he wants it (sex) without using protection. I always take condoms with me, if they do not have, we make them (clients) buy from medical shops. Everybody tells us, advises us to use condoms. [IDI: FSWbetween 35-39 years]

However, periodic economic compulsions and male resistance to condom use were identified as serious constraints on condom use, especially among younger, street based FSWs. Further, the one persistent issue that posed a serious hurdle to adoption of safer sex practices was client violence that often-increased FSWs vulnerability and powerlessness. Street based FSWs in the urban location spoke extensively about their fears and experiences of gender-based violence during the FGD and shared their experiences as depicted in the following quotes:

> He (a paying customer) takes us to a location with a promise to pay us, but then we are exploited…. We become exhausted, we are in pain…[but]They threaten to murder us… If they ask for second time we have to agree… Sometimes they drink liquor and get very violent… if we approach the police, we will be in even greater trouble…

At the same time, there was evidence of FSWs’ varied strategies to prod clients into condom use. As another FGD participant stated:

> Yes, those who have awareness, they use (condoms). Otherwise, we convince them to use. If they don’t listen, I say, ‘I may conceive a child (of course, we are sterilized) and you will have to take care of my child, is it okay?’ At least out of that fear everybody wears a condom. Because nowadays people don’t bother about condoms. They are ready to give more money for not using condoms. In such instances we use this trick. We are unaware about their hidden diseases. We have to take care of our health.

In these purely transactional relationships (Type I), since there was little to no possibility of entering into stable, non-paying, culturally sanctioned partnerships with these men, FSWs’ perceived significant power to initiate condom use in these transactional relationships where they perceived themselves at high risk for HIV/STI. Thus, FSWs were empowered to insist on condom use in these relationships for purposes of HIV and STI risk reduction, particularly because they had no knowledge of such clients’ “hidden diseases” and because they wanted to protect themselves.

##### Emotionally supportive paying partnerships: Type 2

The second type of sexual partnerships that FSWs in this study encountered were clients who return to them regularly. Over half (13) of the in-depth interview participants and many FGD participants described a gradual change from transactional sexual encounters to meaningful relationships over time. As one interviewee described:

> I don’t have sex with every kind of person. Recently I became acquainted with a client, he has very good manners. He now takes care of me really well. He works in a factory. I contact him by telephone whenever I have any problem. Often, he gives money without insisting on sex. He respects me while speaking to me. He calls me whenever he needs me. And we meet frequently. [IDI: FSW between 25-29 years]

Participants’ narrations also highlighted the unique and mutually supportive nature of such (Type 2) relationships, in which there was a gradual change from transactional sexual encounters to meaningful relationships over time. An interviewee in her forties described her relationship with her long-term paying partner in what we categorized as a Type 2 relationship thus:

> Interviewer: Do you have any fear that your partner may leave you? FSW: For me it is ok even if he leaves me. (Laughs)
>
> Interviewer: How did you feel now that he (your partner) has moved far away?
>
> FSW: I feel a lot of pain…But he calls me every day asking if I have eaten enough, and he speaks to the girls also and asks them to take care of me …. that is the love and care he has for me.

Another young FSW in what we describe as a Type 2 relationship spoke at length about the emotional ties she has with one of her clients:

> - He is good person. He will call me once in a while, but he cannot take care of me fully. When I told him about hospitalization of my husband, he scolded me [for not telling him earlier]. He told me that why you had to undergo all this. If you were in trouble I would have come and helped you. …You have given me pleasure at times, I am also a human being, and I will give you money. He gave me money when my husband was hospitalized. Since then, whenever I tell him I have problem, he comes and gives me money and goes back. [IDI: FSW, between 20-24 years]

In these Type 2 relationships, FSW’s partners were perceived as demonstrating care via financial support beyond the sexual transaction, while emphasizing their emotional availability. These kinds of emotionally laden relationships between FSWs and their regular paying partners were further characterized by their mutual affection and trust, combined with calculated assumption of HIV or STI risk. Often, for the respondents, the distinction between paying client and trusted lover became very fluid with such partners. Consequently, participants’ narrations revealed that condom use within such relationships was markedly inconsistent given women’s unwillingness to insist on the same, considering their perceptions regarding their emotional intimacy with such clients. This was despite the clear acknowledgment of the potential risks of non-adoption of safe sex practices, even in these valued Type 2 partnerships. As one participant elucidated:

> My partner never bothers with condoms. If I insist, he refuses, saying that, ‘I’m not having any other affair; I’m not interested in that’. I tell him, ‘I’m an anonymous woman, I met you at a bus stand, are you not bothered whether I’m good [healthy/disease-free] or not?’ He said, ‘I can gauge a person at a glimpse, why should I judge you?’ (Laughs) He does not use condoms. He trusts me very much. Even I trust him completely. Of course, he does not have contact (sexual contact) with any other woman except with me and his wife. I don’t know about his earlier life. I cannot say with certainty that he had no affairs before. But now, he calls me whenever he feels like being in my company…. No I have not used condoms [with him]. Only sometimes…..(condom is used). But, this guy is my guarantee (trusted) companion. [IDI: FSW, between 20-24 years]

In these Type 2 relationships which FSWs interpreted as emotionally supportive, there was the unspoken potential of these paying relationships transitioning into stable non-paying heterosexual partnerships. Resultantly, FSWs’ perception of their bargaining power to use condoms was variable because at times FSWs may have acquiesced to male partners’ desires for non-condom use since these male partners, as stable paying partners, may have had the potential to become stable non-paying partners. Nevertheless, inconsistencies in condom use with Type 2 partners indicated FSWs’ awareness of their heightened HIV /STI risk,

##### Gender-role bound relationships and relationships of convenience: Type 3

Another type of sexual relationship that FSWs in the present study found themselves navigating was with their non-paying sexual partners. These were either spouses or partners with whom they had marriage-like partnerships, and these could involve either cohabiting (socially sanctioned gender-role bound relationships) or non-cohabiting (relationships of convenience). Participants in two FGDs were very vocal about the reasons for needing these kinds of non-paying relationships and the resulting dynamics as the following quotes show:

> For the sake of respect, we should have partners. If we go and ask a house for rent, they will not give, and they will say: ‘You are alone, why should we give?’ They will ask us to bring our husband. Even if we go to some function (social gathering) they will say: ‘She has left her husband; she is a bitch.’ That is one reason madam, another is they (partners) will show love to our kids. So we feel my children are not getting father’s love, he (partner) will give that. I am not getting respect in society because I don’t have husband, I will get that from him.

In the context of these ‘marital-like’ relationships, participants often reported pervasive intimate partner violence and sexual jealousy, which further heightened the women’s sense of gendered vulnerability and their perceptions of lack of personal control and power. Elaborating on their vulnerability in these Type 3 relationships, FG participants stated:

> They are first clients, then they become partners. We don’t get any use out of them. When they are clients, they are different. When they are partners, they change. He will know that we are in this profession. He will not be earning money, or his earning will not be enough for him, as he knows we are earning, he will snatch money. We spend in the thousands for them. He will quarrel, he will beat us. He will come drunk and then beat us. This is very common.
>
> (My partner says) Did I marry you? No. I have kept you (as only a sexual partner). So shut up and stay quiet.

Negotiating condom use in Type 3 relationships was considered largely inappropriate because of the implicit traditional cultural expectation of sexual acquiescence for married Indian women, even within common-law relationships such as described by participants. Hence, despite the awareness of the risk, most respondents expressed frustration at their inability to negotiate condom use with these partners/husbands. As one FGD participant in the urban location chosen stated:

> (Intimate) partners refuse to use condoms. They claim that they are our husbands, though they don’t take any responsibility of a husband. They assume just by having sex, they are a husband to any woman.

In these non-paying relationships FSWs were often in the coveted socially recognized roles and positions of legitimate partners/wives. In exchange for this upgrade in gender capital, FSWs often fully acquiesced to their partner’s demands for non-condom use. These Type 3 relationships were marked by relative lack of power that sex workers experienced and women’s lack of desire to force condom use within socially sanctioned marital/marriage-like often cohabiting relationships. Accordingly, perceived power to negotiate for or to use condoms in these types of relationships was low. Hence, FSWs in Type 3 relationships perceived themselves to be at very high HIV risk.

##### Sexual victimization perpetrators: Type 4

FSWs in the current study recounted instances where they found themselves in dangerous situations where sexual encounters with police, pimps, and gang members were inevitable on an ongoing basis. It was during these situations that respondents experienced the deepest sense of vulnerability and loss of perceived bargaining power and high HIV/STI risk. This was especially true for street based FSWs. Criminalization of sex workers by police was rampant, at times forcing sex workers to go underground. As one participant lamented, “The police observe us, our behavior, our manners, everything. They don’t object our vocation, but they want us to do underground; it should not be a public display…” [IDI: FSW, 20-24 years]. Another sex worker accentuated, “Policemen should give protection to women, even to sex workers. But they are the real criminals. I would like to emphasize this point. They harass us and threaten to arrest us if we don’t satisfy them (sexually)”. [IDI: FSW, between 30-34 years]

Along with police, FSW in our sample reported being subjected to sexual assault and other forms of violence by gang members (rowdies) and by law enforcement officers. A FGD participant described these experiences such:

> Rowdies also torture us. I may believe a customer and go with him to a farm. I will find an additional 3 or 4 rowdies waiting for me [to have sex with them also]. I will be caught and raped by all the others…

Here is an example of an FSW whose statement demonstrates the way she actively navigates different types of sexual partnerships and HIV risk as depicted in Figure 1: Yes, for those types of clients I keep a stock (of condoms). Also, I tell them, “Use it, otherwise I will not go with you…” But for the regular partner, he wanted to marry. He was ready to give me a thaali (gold chain symbolic of a legally married woman), but I refused. I said: “You are my lover. I cannot have more than one husband.” He told me he will build a house for me and take care of me….but I don’t want it. We can meet, whatever our needs are we can satisfy that and come back. Then there is the partner who I live with…he tortures me a lot. He is very controlling, says: “Don’t go here, don’t go there. Don’t meet him..” Many a time he has beaten me severely…I have had injuries to my head. Even my husband (who is dead now) never tortured me so much And then, I have to deal with the rowdies who often come in group, they torture more and don’t pay money. They insist on having sex without condom and go away without paying. They threaten me saying they will inform the police. We talk of police but the police themselves would come and ask for girls (for sex). They will ask: “Why one should wear it (condom)?” They offer more money and say: “Why don’t you do it (sex) without condom?

[IDI: FSW between30-34 years]

## Discussion

The present study is among the few that have responded to previous calls for research to document the finer distinctions of emotion-laden romantic relationships and meaningful sexual partnerships for FSWs (Deering et al., 2011). Findings from this phenomenological exploration contribute to the extant literature on FSWs’ journeys through fluid sexual partnerships and their assessment of risk and condom use negotiation. Although previous studies (Bharat et al., 2013) have reported low condom use within FSWs’ intimate relationships, they documented the concurrent multiple relationships that sex workers navigate as separate and mutually exclusive. In the present study, we demonstrated that while some partnerships appeared to remain in the realm of *commercial* transactions, others that began that way morphed into one that exemplified *emotional intimacy and romance*. Still other partnerships occupied the space of *relationships of convenience*. Significantly, women appeared to simultaneously juggle and straddle issues of mere exchange, intimacy, convenience, and gender role expectations and overt power with various paying and non-paying partners like that found in prior research with sex workers in Mumbai city in India (Karandikar & Prospero, 2010). Also, ideas from past studies in other parts of the global South on transactional sex and the fluidity of sex work (Decoteau, 2016; Hoang, 2011; Mojola, 2015; Swidler, et al., 2007) have not been applied as much in the Indian context where research has largely viewed clients through a narrow lens. Our study adds to existing literature by highlighting the nuances of the morphing of transactional exchanges into emotionally laden romantic relationships with clients and the fluid line between ‘sex work’ and ‘relationship.’

Participant narrations revealed that their entry into sex work often stemmed out of their proactive efforts to escape abusive and unfulfilling heterosexual marriages/partnerships to support their families, enabled by their female social networks. This finding is important because it reflects the Gendered Vulnerability described earlier. The current study sample was like what is characterized by Stoebenau and colleagues (2009) as “low sex work,” consisting of individuals who were “usually very poor” and who were often involved in fluid relationships in efforts to secure socially desirable stable male partners as well as economic stability. Nevertheless, socioeconomically disadvantaged FSWs in our study enacted agency in their striving for some semblance of social inclusion within their highly constrained life circumstances. Through navigating relationship fluidity, which at times resulted in stable heterosexual partnerships, these Indian FSWs made the most with the little to no social capital they possessed. Prior studies conducted with street and home-based FSWs in Karnataka state and Chennai city in India have shown that even though involvement in heterosexual partnerships rendered these women vulnerable to possible HIV infection (through non-condom use) as well to intimate partner victimization, the gendered role as wife/spouse to a non-paying male partner is valued within Indian society and perceived as less desirable than a non-wife/spouse status for women (Deering et al., 2015; Panchanadeswaran et al., 2008). The findings from the study extend the understanding of these issues by highlighting women’s trajectories of HIV risk tolerance within the various relationships, particularly within those relationships that they evaluate as meaningful based on their perceptions of intrinsic rewards, e.g., being available as an emotional connection for some paying customers in addition to receiving monetary incentives. Importantly, this study emphasizes the dynamic and fluid nature of not only sexual partnerships, but also fluidity of risk that shifts both within and between partnerships based on ascribed meanings to the partnerships. Results from this study also challenge dominant discourses that focus on sex workers primarily as vectors of disease and underscores not only the complexity of their sexual partnerships but demonstrates that sex workers’ relationships with paying clients maybe among the least dangerous.

Consistent with prior studies elsewhere and in India (Deering et al. 2011; Ghose et al., 2011; Rosenthal & Oanh, 2006; Warr & Pyett, 1999), FSWs in the current study reported greater insistence and willingness to negotiate for condom use with one time paying clients but not with those they perceive as intimate partners. Women’s efforts to demarcate their personal and professional spheres thus consciously were like that highlighted in earlier research (Brewis & Linstead, 2000) where power and intimacy varied. However, the current study showed that the reticence to use condoms or to negotiate for condom use posed additional hurdles when dealing with *partners in relationships of convenience* who served as crucial co-actors in FSWs’ quest to maintain a semblance of the sanctioned heterosexual familial relationships that reduce societal stigma, overt discrimination and violence from gangs, landlords, and police officers. Additionally, when FSWs in our study perceived lack of judgment by non-paying partners as trust, they did not insist on condom use pointing to the major role of personal assessments of power, control, and emotional attachments while balancing HIV risk.

Findings from the current study also highlighted how the location of sex work, i.e. home/street-based sex work played an important role in FSWs’ condom use decisions. It appeared that FSWs’ personal networks and relationships with lodge owners offered them vital protection and security when things went awry with hostile clients. On the other hand, when streets/fields/farms were the site of sex work, this placed women at significantly higher risk for threats and severe violence by exploitative clients; thereby increasing FSWs’ vulnerability and higher resistance to negotiate condom use.

Some limitations of the study bear mention. Given that sex workers in India are a hidden population due to their stigmatized status, non-probability sampling techniques were used to recruit a modest sample of 60 FSWs which nevertheless permitted rich exchanges of ideas and dynamic interaction among the key stakeholders (in our FGD) as well as detailed dialogue in the semi-structured in-depth interviews (Morse, 2000). However, the self-reported data can lead to social desirability bias and be affected by participants’ capacity to correctly recall the events and relationships they described in this study (Bergen & Labonté, 2020). Also, despite this study’s important contributions, we did not unpack the differential experiences of FSWs across geographical locations, caste, and age categories. Future studies would benefit from using an intersectional lens to examine the experiences of the diverse population of FSWs in India. Also, these data were collected prior to the COVID-19 pandemic and hence may not reflect any significant shifts in the sex work terrain that may have ensued post 2020, which would benefit from exploration in future studies. Finally, we cannot generalize the findings beyond the Karnataka state in India where the study was conducted.

## Implications

However, these limitations notwithstanding, findings from the current study have several important implications. Our findings showed that the fluidity of sexual partnerships makes condom negotiation and use considerably complex for FSWs in India. This underscores the need for interventions that consider the continuum of risk, the dynamic nature of risks as relationships shifts from paying to non-paying and so forth, and the subjective appraisal of risk that dictate FSWs’ decision making. Thus, social workers and public health practitioners engaging in HIV prevention efforts need to recognize that HIV knowledge is a necessary but not sufficient condition that determined ability and willingness to negotiate condom use. Importantly, it is critical to comprehend the stigma surrounding sex work and sex workers in India that poses a formidable barrier to consistent condom use which further highlights the urgent need to recognize sex work as legitimate work (Deering et al. 2011; Panchanadeswaran et al., 2016; Vijayakumar et al., 2015a, 2015b). The dynamic, non-linear, and fluid nature of relationships as well as risk assessments need to be understood with the broader sexual culture within which these relationships are embedded. That is salient, given that researchers have stressed the need to acknowledge the role of non-commercial sexual partners as key bridge populations in the spread of infections in India (Deering et al., 2011). Significantly, the dangers posed by even subtle broaching of condom use within certain sexual encounters cannot be underestimated. Prevention efforts would need to account for FSWs’ experiences of violence as a key risk factor not only from intimate partners but street violence and sexual assault from gangs and state actors such as police personnel.

Explicit focus on both individual and collective empowerment through HIV programs and sex worker unions can result in enduring advantages for FSWs, including reduced vulnerability and increased autonomy (Blanchard et al., 2013). The inclusion of men consistently in public health campaigns is vital as is normalizing men’s proactive use of condoms. A nuanced understanding of FSWs’ complex relationship contexts and the tradeoffs are vital while designing effective interventions for HIV prevention. Insights around behavioral norms and decision making (in the form of relationship fluidity) could be critical in the planning and implementation of HIV-prevention programs among FSWs. It is vital to strongly counter policies such as the proposed draconian anti-trafficking bill (The Trafficking in Persons Prevention, Care and Rehabilitation Bill, 2021) that problematically combines human trafficking and sex work and therefore, significantly increases the chances of violent victimization of sex workers by state actors such as the police (Bhattacharya, 2021). It is critical to not view these women as victims who need rescuing, but individuals that have exercised their agency in choosing their line of work. Structural interventions to decriminalize sex work should also provide opportunities for greater access to gendered social viability through mechanisms such as community organization, outreach, peer leadership and building solidarity with other allied networks (Shannon et al., 2015) within a heteropatriarchal society that stigmatizes and criminalizes sex workers. Additionally, accounting for cultural milieus that often determine women’s choices as well as decision-making is vital. Finally, it is critical to develop an empathic understanding of sex workers’ lives and use it in the design of programs and other interventions (Tavory & Poulin, 2012).

## Data Availability

All data produced in the present study are available upon reasonable request to the authors.

## Acknowledgements

This research was supported by a Faculty Development Grant from Adelphi University to the first author (PI, Subadra Panchanadeswaran; Co-I, Sel J. Hwahng). The authors would like to gratefully acknowledge Dr. Gowri Vijayakumar, Ms. Elizabeth Chemick, Ms. Justy John, Ms. Joanna Barberii, & Ms. Natalie Brooks-Wilson, and Ms. Seungju Lee for their help with this paper.

## References

Ahmed, A., & Seshu, M. (2012). “We Have the Right Not to Be ‘Rescued”: When Anti-Trafficking Programmes Undermine the Health and Well-Being of Sex Workers. Anti-Trafficking Review, 1(103), 149–68.

Azhar, S., Dasgupta, S., Sinha, S., & Karandikar, S. (2020). Diversity in sex work in India: Challenging stereotypes regarding sex workers. Sexuality & Culture, 24, 1774–1797. https://doi.org/10.1007/s12119-020-09719-3

Bergen, N., & Labonté, R. (2020). “Everything Is perfect, and we have no problems”: Detecting and limiting social desirability bias in qualitative research. Qualitative Health Research 30(5), 783–792. http://globalhealthequity.ca/wpcontent/uploads/2020/07/2020_QHR_Social-desirability-bias-1.pdf

Bharat, S., Mahapatra, B., Roy, S., & Saggurti, N. (2013). Are female sex workers able to negotiate condom use with male clients? The case of mobile FSWs in four high HIV prevalence states of India. PloS one, 8(6), e68043.

Bhattacharya, S. (2021, July 12). Why sex workers’ organisations aren’t pleased with the Draft Anti-Trafficking Bill. The Wire. https://thewire.in/rights/draft-anti-trafficking-bill-sex-workers

Blanchard, A. K., Mohan, H. L., Shahmanesh, M., Prakash, R., Isac, S., Ramesh, B. M., … & Blanchard, J. F. (2013). Community mobilization, empowerment and HIV prevention among female sex workers in south India. BMC Public Health, 13(1), 1–13.

Brewis, J., & Linstead, S. (2000). ‘The worst is in the screwing’ (1): Consumption and the management of identity in sex work. Gender, Work, and Organization, 7(2), 84–97. https://doi.org/10.1111/1468-0432.00096

Campbell, J. L., Quincy, C., Osserman, J., & Pedersen, O. K. (2013). Coding in-depth semistructured interviews: Problems of unitization and intercoder reliability and agreement. Sociological Methods & Research, 42(3), 294–320.

Creswell, J. W. (1998). Qualitative inquiry and research design: Choosing among five traditions. Thousand Oaks, Calif: Sage Publications.

Decoteau, C.L. (2016). ‘You Can’t Eat Love’: ‘Getting by’ in South Africa’s Informal Sexual Economy. American Journal of Cultural Sociology, 4 (3), 289–322.

Deering, K. N., Bhattacharjee, P., Bradley, J., Moses, S. S., Shannon, K., Shaw, S. Y., … & Rajaram, S. (2011). Condom use within non-commercial partnerships of female sex workers in southern India. BMC public health, 11(6), S11.

Deering, K. N., Shaw, S. Y., Thompson, L. H., Ramanaik, S., Raghavendra, T., Doddamane, M., … & Lorway, R. (2015). Fertility intentions, power relations and condom use within intimate and other non-paying partnerships of women in sex work in Bagalkot District, South India. AIDS care, 27(10), 1241–1249.

Dutton, M. A., Goodman, L., & Schmidt, R. J. (2006). Development and Validation of a Coercive Control Measure for Intimate Partner Violence: Final Technical Report. Retrieved from https://www.ncjrs.gov/pdffiles1/nij/grants/214438.pdf

Drigotas, S.M., & Rusbult, C.E. (1992). Should I stay or should I go? A dependence model of breakups. Journal of Personality and Social Psychology, 62, 62–87. https://doi.apa.org/doiLanding?doi=10.1037%2F0022-3514.62.1.62

Erausquin, J. T., Reed, E., & Blankenship, K. M. (2011). Police-related experiences and HIV risk among female sex workers in Andhra Pradesh, India. Journal of Infectious Diseases, 204(Suppl_5), S1223–S1228.

Fehrenbacher, A. E., Chowdhury, D., Ghose, T., & Swendeman, D. (2016). Consistent condom use by female sex workers in Kolkata, India: Testing theories of economic insecurity, behavior change, life course vulnerability and empowerment. AIDS and Behavior, 20(10), 2332–2345. doi:10.1007/s10461-016-1412-z

Fitzgerald-Husek, A., Martiniuk, A. L., Hinchcliff, R., Aochamus, C. E., & Lee, R. B. (2011). “I do what I have to do to survive”: An investigation into the perceptions, experiences and economic considerations of women engaged in sex work in Northern Namibia. BMC women’s health, 11(1), 35. doi: 10.1186/1472-6874-11-35

Ghose, T., Swendeman, D. T., & George, S. M. (2011). The role of brothels in reducing HIV risk in Sonagachi, India. Qualitative Health Research, 21(5), 587–600.

Glaser, B. G., & Strauss, A. L. (1967). The discovery of grounded theory: strategies for qualitative theory. New Brunswick: Aldine Transaction.

Hoang, K.K.(2011). ‘She’s Not a Low-Class Dirty Girl!’: Sex Work in Ho Chi Minh City, Vietnam. Journal of Contemporary Ethnography, 4 (4), 367–96.

Hui, N. (2017). Bargaining power and indicators of well-being among brothel-based sex workers in India. Feminist Economics, 23 (3), 49–76.

Jaggar, A. M. (2009). Transnational cycles of gendered vulnerability: A prologue to a theory of global gender. Philosophical Topics, 37(2), 33–52. https://www.jstor.org/stable/43154555

Jie, W., Xiaolan, Z., Ciyong, L., Moyer, E., Hui, W., Lingyao, H., & Xueqing, D. (2012). A qualitative exploration of barriers to condom use among female sex workers in China. PloS one, 7(10), e46786. doi:10.1371/journal.pone.0046786

Joshi, M., Rahill, G. J., Lescano, C., & Jean, F. (2014). Language of sexual violence in Haiti: perceptions of victims, community-level workers, and health care providers. Journal of health care for the poor and underserved, 25(4), 1623–1640. doi: 10.1353/hpu.2014.0172.

Kaiwar, A., & Gothoskar, S. (2014). “Who Says We Do Not Work?” Economic and Political Weekly 49 (46), 54–61.

Karandikar, S., & Prospero, M. (2010). From Client to pimp: Male violence against female sex wokers. Journal of Interpersonal Violence, 25(2), 257–273. https://doi-org.libproxy.adelphi.edu/10.1177%2F0886260509334393

Karandikar, S., Casassa, K., Knight, L., Espana, M., & Kagotho, N. (2022). “I am almost a breadwinner for my family:” Exploring the manifestation of agency in sex workers’ personal and professional contexts. Affilia: Feminist Inquiry in Social Work, 37(1), 26–41. https://doi-org.libproxy.adelphi.edu/10.1177%2F08861099211022717

Kotiswaran, P. (2001). Preparing for Civil Disobedience: Indian Sex Workers and the Law, 21 B.C. Third World L.J. 161. https://lawdigitalcommons.bc.edu/twlj/vol21/iss2/1

Kotiswaran, P. (2011). Dangerous sex, invisible labor: Sex work and the law in India. Princeton, NJ: Princeton University Press.

Mojola, A. (2015). Material Girls and Material Love: Consuming Femininity and the Contradictions of Post-Girl Power among Kenyan Schoolgirls. Continuum, 29 (2), 218–29.

Morse, J. M. (2000). Determining sample size. Qualitative Health Research, 10(1):3–5. https://doi.org/10.1177/104973200129118183

Muhr, T. (2013). ATLAS.ti 7.0 user’s manual and reference (Vol. 7). Berlin: Scientific Software Development GmbH.

Murray, L., Moreno, L., Rosario, S., Ellen, J., Sweat, M., & Kerrigan, D. (2007). The relationship intimacy in consistent condom use among female sex workers and their regular paying partners in the Dominican Republic. AIDS and Behavior, 11(3), 463–470.

National AIDS Control Organization & ICMR-National Institute of Medical Statistics (2020). India HIV Estimates 2019: Report. New Delhi: NACO, Ministry of Health and Family Welfare, Government of India. Retrieved on September 19, 2021 from http://naco.gov.in/sites/default/files/Estimation%20Report%202019.pdf.

Ngugi, E., Benoit, C., Hallgrimsdottir, H., Jansson, M., & Roth, E. A. (2012). Partners and clients of female sex workers in an informal urban settlement in Nairobi, Kenya. Culture, health & sexuality, 14(1), 17–30.

Odinokova, V., Rusakova, M., Urada, L. A., Silverman, J. G., & Raj, A. (2014). Police sexual coercion and its association with risky sex work and substance use behaviors among female sex workers in St. Petersburg and Orenburg, Russia. International Journal of Drug Policy, 25(1), 96–104.

Panchanadeswaran, S., Johnson, S. C., Sivaram, S., Srikrishnan, A. K., Latkin, C., Bentley, M. E., … & Celentano, D. (2008). Intimate partner violence is as important as client violence in increasing street-based female sex workers’ vulnerability to HIV in India. International Journal of Drug Policy, 19(2), 106–112.

Panchanadeswaran, S., Johnson, S. C., Sivaram, S., Srikrishnan, A. K., Zelaya, C., Solomon, S., … & Celentano, D. (2010). A descriptive profile of abused female sex workers in India. Journal of health, population, and nutrition, 28(3), 211.

Panchanadeswaran, S., Vijayakumar, G., Chacko, S., & Bhanot, A. (2016). Unionizing Sex Workers: The Karnataka Experience. In Special Issue: Problematizing Prostitution: Critical Research and Scholarship (pp. 139-156). Emerald Group Publishing Limited.

Panchanadeswaran, S., Unnithan, A.M., Chacko, S., Brazda, M., & Kuruppu, S. (2017). What’s technology got to do with it? Exploring the impact of mobile phones on female sex workers’ lives and livelihood in India. Gender, Technology & Development, 21(1-2), 152–167. https://doi.org/10.1080/09718524.2017.1385318.

Patterson, T. L., Mausbach, B., Lozada, R., Staines-Orozco, H., Semple, S. J., Fraga-Vallejo, M., … & Martinez, G. (2008). Efficacy of a brief behavioral intervention to promote condom use among female sex workers in Tijuana and Ciudad Juarez, Mexico. American journal of public health, 98(11), 2051–2057.

Pulerwitz, J., Gortmaker, S.L., & DeJong, W. (2000). Measuring relationship power in HIV/STD research. Sex Roles, 42, 637-660. Doi:10.1023/A:1007051506972

Rosenthal, D., & Oanh, T. T. K. (2006). Listening to female sex workers in Vietnam: influences on safe-sex practices with clients and partners. Sexual health, 3(1), 21–32.

Shahmanesh, M., Patel, V., Mabey, D., & Cowan, F. (2008). Effectiveness of interventions for the prevention of HIV and other sexually transmitted infections in female sex workers in resource poor setting: a systematic review. Tropical Medicine & International Health, 13(5), 659–679.

Shannon, K., Strathdee, S. A., Goldenberg, S. M., Duff, P., Mwangi, P., Rusakova, M., … & Boily, M. C. (2015). Global epidemiology of HIV among female sex workers: influence of structural determinants. The Lancet, 385(9962), 55–71. DOI:10.1016/S0140-6736(14)60931-4

Stoebenau, K., Hindin, M. J., Nathanson, C. A., Rakotoarison, P. G., & Razafintsalama, V. (2009). “… But then he became my sip”: the implications of relationship fluidity for condom use among women sex workers in Antananarivo, Madagascar. American journal of public health, 99(5), 811–819.

Summer, A., Guendelman, S., Kestler, E., & Walker, D. (2017). Professional midwifery in Guatemala: A qualitative exploration of perceptions, attitudes and expectations among stakeholders. Social Science & Medicine, 184, 99–107.

Swidler, A., & Cotts Watkins. (2007). Ties of Dependence: AIDS and Transactional Sex in Rural Malawi. Studies in Family Planning, 147–62.

Tavory, I., & Poulin, M. (2012). Sex work and the construction of intimacies: meanings and work pragmatics in rural Malawi. Theory and society, 41(3), 211–231.

UNAIDS (2014). The Gap Report 2014. Retrieved on January 26, 2022 from https://www.unaids.org/sites/default/files/media_asset/06_Sexworkers.pdf

VanderDrift, L.E., Agnew, C.R., Harvey, S.M., & Warren, J.T. (2013). Whose intentions predict? Power over condom use within heterosexual dyads. Health Psychology, 32 (10), 1038–1046.

Vijayakumar, G., Chacko, S., & Panchanadeswaran, S. (2015a). Sex workers join the labor movement in India. New Labor Forum, 24(2), 90–96.

Vijayakumar, G., Chacko, S., & Panchanadeswaran, S. (2015b). Sex workers and the informal labor movement in India. Global Labor Journal, 6(1), 79–96.

Vijayakumar, G. (2021). At Risk: Indian sexual politics and the global AIDS crisis. Stanford, CA: Stanford University Press.

Warr, D. J., & Pyett, P. M. (1999). Difficult relations: sex work, love and intimacy. Sociology of Health & Illness, 21(3), 290–309.

